# Dementia and hearing-aid use: a two-way street

**DOI:** 10.1101/2021.10.25.21265439

**Authors:** Graham Naylor, Lauren Dillard, Martin Orrell, Blossom Stephan, Oliver Zobay, Gabrielle Saunders

## Abstract

**Objectives:** Hearing-aid use may reduce risk of dementia, but cognitive impairment makes hearing-aid use more challenging. These two causal pathways may both manifest as an association between reduced hearing-aid use and incident dementia. This study examined the effects of each pathway separately, through a unique combination of longitudinal data regarding hearing, general health, dementia diagnoses and continuity of HA use.

**Methods:** Longitudinal health records data from 380,794 Veterans who obtained hearing aids from the US Veterans Affairs healthcare system were analysed. Analysis 1 (n=72,180) used logistic regression to model the likelihood of a dementia diagnosis at 3 year 6 months (3y6m) to 5 years post hearing-aid fitting for patients previously free of dementia and mild cognitive impairment (MCI). Analysis 2 (n=350,918) modelled the likelihood of being a persistent hearing aid user at 3y2m after fitting, contrasting sub-groups with differing levels of cognitive function at the time of fitting. Models controlled for relevant predictors available in the dataset.

**Results:** In analysis 1, the adjusted OR for incident dementia was 0.73 (ci 0.66-0.81) for persistent (vs. non persistent) hearing-aid users. In analysis 2, the adjusted OR for hearing-aid use persistence was 0.46 (ci 0.43-0.48) in those with pre-existing dementia (vs. those remaining free of MCI and dementia).

**Conclusion:** The results indicate substantial effects from both causal pathways. Research studying protective effects of hearing-aid use against dementia needs to account for this. Clinically, hearing devices and hearing care processes must be accessible and usable for all, regardless of their level of cognitive function.

## Introduction

Modifiable risk factors for dementia include untreated mid-life hearing loss, and it has been estimated that 8% of dementia cases globally are attributable to this factor (Livingston et al., 2020). Proposed mechanisms responsible for the relationship between hearing loss and the development of dementia include (1) common underlying pathology (probably vascular), (2) impoverished input affecting brain structure and function, (3) cognitive resources over-occupied in listening unavailable for higher functions, and (4) interaction between auditory function and dementia pathology (Griffiths et al., 2020). These mechanisms are not mutually exclusive.

The primary treatment for hearing loss is the fitting of hearing aids (HA). One may hypothesize that treatment with HA will alleviate or decelerate cognitive decline (forward causal path: HA use -> function). However, the outcome will depend on which of the above mechanisms is dominant, and when the intervention takes place relative to the onset of hearing loss.

Meanwhile, since cognitive decline is associated with difficulty in maintaining instrumental activities of daily living (Pérès et al., 2008), it is also logical to hypothesize that adherence to recommended HA treatment is lower in those with cognitive decline (reverse causal path: function -> HA use). Ageing is associated with increased risk of cognitive decline, dementia and hearing loss. Thus, if HAs are to slow cognitive decline, they must be used and usable by people whose cognitive function may already be impaired.

We begin by reviewing evidence relating to the two causal pathways outlined above.

In a large study (n=114,862) of individuals aged >66 years just diagnosed with hearing loss, Mahmoudi et al. (2019) found that those who acquired HAs had a decreased likelihood of 3-year incident Alzheimer’s disease (AD)/dementia compared to those who did not. This is indicative of a causal association (HA use -> function), although data about HA use over time were absent.

Dawes et al. (2015) analysed 16-year longitudinal data from 666 participants with hearing loss, and found no differences between HA users and non-users in cognitive outcomes, even when accounting for changes in HA usage over time, suggesting that HA use does little to prevent or slow cognitive decline.

Three other studies (Amieva et al., 2018; Ray et al., 2018; Maharani et al., 2018) provide some evidence of support for the forward pathway. All three studies concluded that HA users show less cognitive decline over time than non-users, but in our view each has serious shortcomings with respect to whether HA use provided a protective effect against dementia. In the study by Amieva et al. (2018), hearing problems and HA use were only assessed at baseline, which was 25 years prior to outcome measurement. Ray et al. (2018) carried out a cross-sectional analysis of data from one wave of a longitudinal study, hence causality cannot be inferred. The statistical model of Maharani et al. (2018) indicated that the rate of cognitive decline after HA fitting was indeed slower than before fitting, but also that cognitive function declined abruptly at the time of HA fitting. Furthermore, the analysis assumed that HA use continued indefinitely.

It deserves mention that the influential Lancet Commission report (Livingston et al., 2020) citing these same three studies, states “The long follow-up times in these prospective studies suggest hearing aid use is protective, rather than the possibility that those developing dementia are less likely to use hearing aids.” However, none of these studies controlled for HA use persistence, and one is cross-sectional, therefore we do not find Livingston et al.’s rejection of reverse causation to be robust. Indeed, Livingston et al. explicitly point out elsewhere in their report that in order to have any helpful effect, HAs have to be usable by the patients concerned.

We now turn to the reverse causal pathway (function -> HA use), for which three studies indirectly provide support. Cohen-Mansfield and Taylor (2002) interviewed two hundred and seventy-nine nursing home residents and 51 nursing staff about barriers to HA use amongst residents, many of whom had cognitive decline. Reported problems included intolerance of devices and difficulty using and maintaining HAs. These occurred with higher rates than in cognitively intact populations, and HA use rates were substantially lower. Deal et al. (2015) analysed data from 253 older adults obtained over 23 years. Among 85 participants with mild/severe HL, those who reported using HAs showed a significantly slower rate of cognitive decline over the preceding 20-year period, even after controlling for a range of confounds. The authors interpreted this as “support [for] future research on whether HA treatment may reduce risk of cognitive decline.” However, HA use was only assessed at the end of the study, so if anything, the results point to the reverse direction of causality. In the third study, Gregory et al. (2020) carried out qualitative interviews with sixteen HA owners with MCI or mild AD were conducted to explore experiences of HA use. Four themes emerged: (1) memory problems reduce use; (2) practical issues reduce use; (3) most participants found HAs to be beneficial; but (4) some show ambivalence about the need for HAs. While these themes may not be unique to those with cognitive impairment (Bertoli et al., 2009; Fisher et al., 2011; Gopinath et al., 2011), theme 1 is very likely to be accentuated in this population, and possibly theme 2 also.

Therefore, in this paper we attempt to minimize the limitations of either cross-sectional or longitudinal data in which HA use and/or cognitive status are only assessed at a single time point, in order to separately estimate the magnitude of the effects of the two potential causal pathways. Specifically, using data from the Veterans Affairs (VA) electronic health record (EHR) system of Veterans who had received hearing aids through the VA system, we investigate the hypotheses:

1. Persistent hearing aid use in cognitively intact Veterans aged 60+ will be longitudinally associated with reduced risk of incident dementia,
2. Pre-existing dementia in Veterans aged 60+ will be longitudinally associated with reduced hearing aid use persistence.

## Materials and methods

This work was approved by the Institutional Review Board and the Research and Development Committee of the VA Portland Health Care System (Study #03566), Data Access Request Tracker (tracking number 2014-11-066-D-A04), and VA Patient Care Services.

### Population

Data were obtained from the VA EHR system. Within the VA healthcare system, eligible Veterans are provided with HAs and batteries (as needed on request) free of charge, each of which are documented in the EHR system. As described in detail previously (Saunders et al., 2021), the initial dataset comprised all patients in the VA EHR system with a HA order between 2012/04/01 and 2014/10/31 (N=731,213). The following data were available: all diagnostic (International Classification of Diseases; ICD) and procedural codes for the period 2007/01/01 (or earliest occurrence thereafter) until 2017/12/31; HA order data between 2012/04/01 to 2014/10/31; HA battery order data for 2012/04/01-2017/12/31.

Patients meeting the following criteria were included: (1) Surviving until 2017/31/12; (2) a single HA order and HA fitting, between 2012/04/01 and 2014/10/31, with at most 180 days between order date and fitting date; (3) ≥60 years of age at time of HA fitting; and (4) available audiometric data. After applying these criteria, the sample size was n=380,794. Of these, 98.9% were male.

The sample was filtered by additional temporal and diagnostic criteria to generate sub-groups of patients for the specific purposes of testing the above hypotheses 1 and 2 in analyses 1 and 2 respectively (see Figure 1).

**Figure 1:**
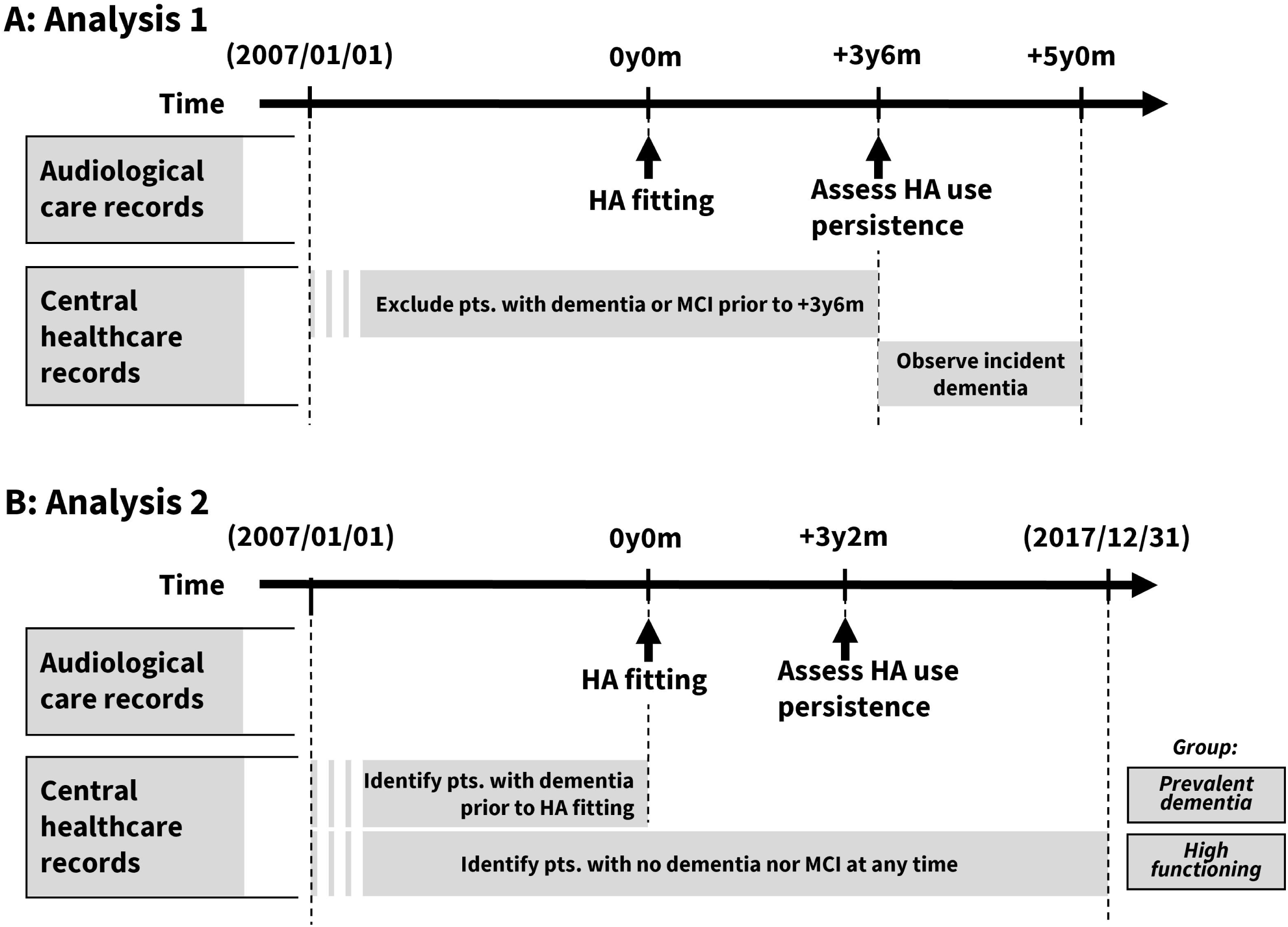
The temporal arrangement of the data used to determine inclusion and extract variables for analysis 1 (A, upper) and analysis 2 (B, lower). See text for further explanation.

### Analysis 1: Does persistent HA use reduce the risk of incident dementia?

#### Sample

We included at baseline only patients free of diagnoses of dementia (see Supplementary Data S1 for ICD-9 and ICD-10 codesets) or MCI (see Supplementary Data S2 for ICD-9 and ICD-10 codesets) up to the start of the time window for monitoring incident dementia (here, 3y6m after HA fitting). The analysis is complicated by the ICD system switch from version 9 to 10 on 2015/10/01. In order that all monitoring of incident dementia was based on only one system, we constrained the incidence time window to begin no earlier than 2015/10/01. To maintain a reasonable balance between the lengths of the HA use persistence (and prevalent conditions) window, the length of the incident dementia window, and the available N, the patient sample for analysis 1 was restricted to those with HA fittings up to 2012/12/31. Applying these criteria, the sample size for this analysis was n=72,180.

#### Variables and analysis

##### Incident Dementia

Incident dementia was recorded if at least one diagnosis from the ICD-10 codeset (Supplementary Data S1) occurred between 3y6m and 5 years after HA fitting.

##### Covariates

Binary variables were constructed for the presence of each of the following dementia risk factors, based on diagnoses recorded up to 3y6m post HA fitting: obesity, stroke, diabetes, depression, bipolar disorder and hypertension. See Supplementary Data S3 for the ICD-9 and ICD-10 codesets used. Additional variables included age (years) at time of HA fitting and pure-tone average hearing threshold (PTA, dB HL) computed by averaging thresholds at 0.5, 1, 2 and 4 kHz for both ears.

A binary proxy variable indicating whether HA use remained persistent at 3y6m post HA fitting was determined for each patient based on their history of HA battery orders, according to our previously published derivation (Zobay et al., 2021). In brief, a HA user is deemed to remain persistent at time *t* if they have ordered a pack of batteries within the 18-month period leading up to *t*.

Logistic regression was used to model the likelihood of incident dementia 3y6m to 5 years post HA fitting. The analysis controlled for HA use persistence at 3y6m post fitting (binary); age (to second order); PTA; and diagnosis of each of obesity, stroke, diabetes, depression, bipolar disorder, and hypertension.

### Analysis 2: Does cognitive function at time of HA fitting predict subsequent HA use persistence?

#### Sample

To investigate the impact of cognitive function on HA use we included only those patients whose cognitive status was clearly defined by diagnostic codes and remained unchanged from HA fitting onwards:

- a ‘prevalent dementia’ group (N=6,418) of patients whose EHRs included at least one diagnosis of age-related dementia (ICD-9 codeset, Supplementary Data S1) prior to HA fitting, and
- a ‘normal cognition’ group (N=344,500) of patients whose EHRs contained no diagnoses of dementia or MCI (ICD-9 or ICD-10 codesets, Supplementary Data S1 and S2) whatsoever, up to 2017/12/31.

For both groups, a clearance period criterion was applied, such that patients were only included if their first recorded VA outpatient visit was at least 2 years before HA fitting.

#### Variables and analysis

Logistic regression was used to model the likelihood of HA use persistence at 3y2m post-fitting (determined in the same way as Analysis 1), this being the most distal post-fitting time for which all patients could be included equitably (due to the dataset cut-off at 2017/12/31) while maximizing the likelihood that a patient who is going to discontinue HA use will have already done so.

The model included group (prevalent dementia vs. normal cognition), and additional variables known to be strongly associated with HA use persistence (Saunders et al., 2021): Multimorbidity index using the Chronic Condition Indicator (CCI; Healthcare Cost and Utilization Project, 2016) for the 12-month period prior to HA order, after removal of codes for hearing loss and mental health to avoid overlap with primary variables; Age (years) at time of HA fitting (to second order); New HA recipient vs. Experienced HA user (binary variable provided in the EHR (Saunders et al., 2021); PTA (dB HL).

## Results

### Analysis 1: Does persistent HA use reduce the risk of incident dementia?

Table 1 presents the OR, corresponding 95% CI and p-values for predictors in the logistic regression model of analysis 1. Figure 2 shows the unadjusted and adjusted dementia incidence estimates, grouped by HA use persistence and age decade.

**Table 1:**
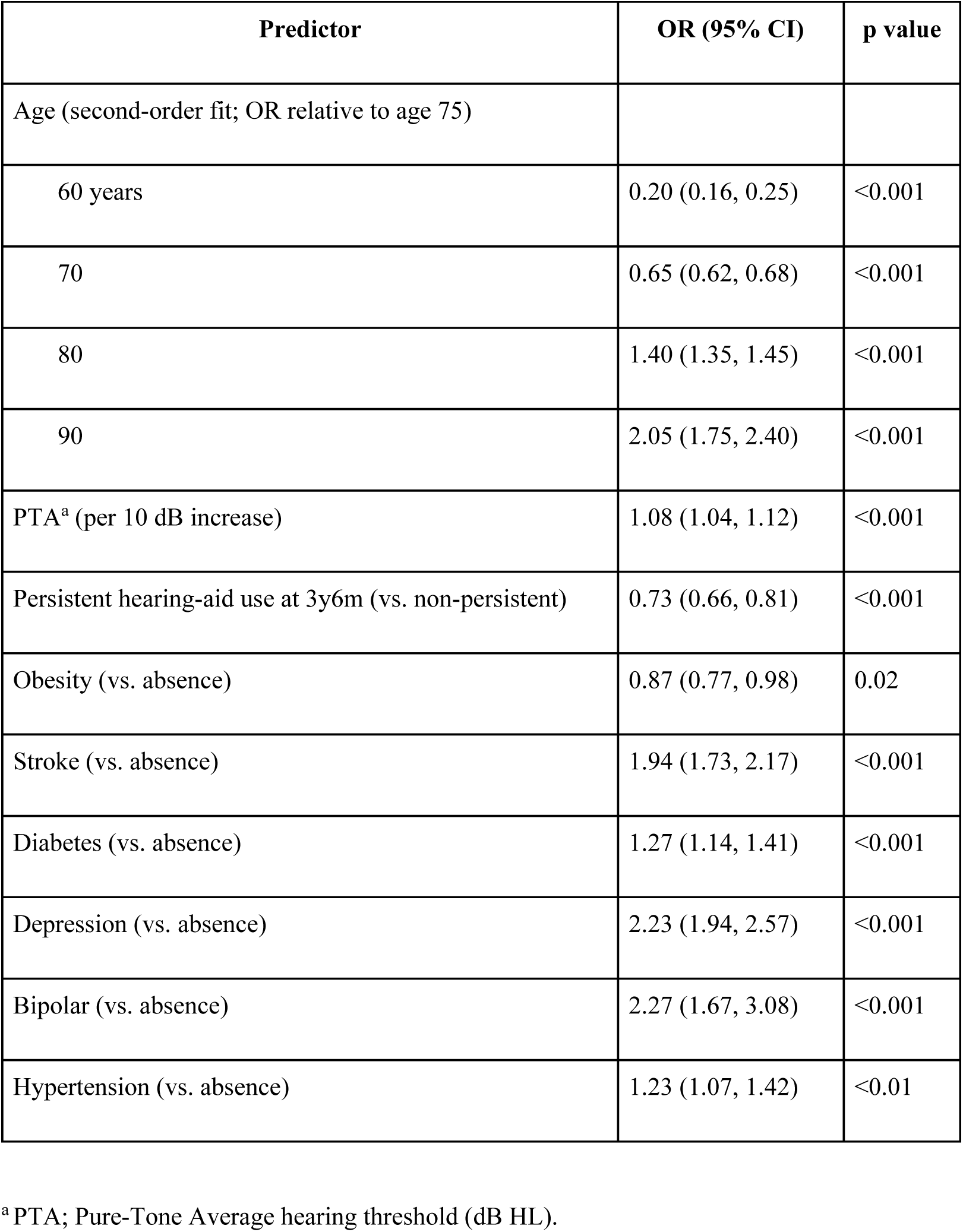
Logistic regression model for Analysis 1; likelihood of dementia diagnosis between 3y6m and 5y after hearing-aid fitting in patients free of dementia and MCI up to 3y6m.

**Figure 2:**
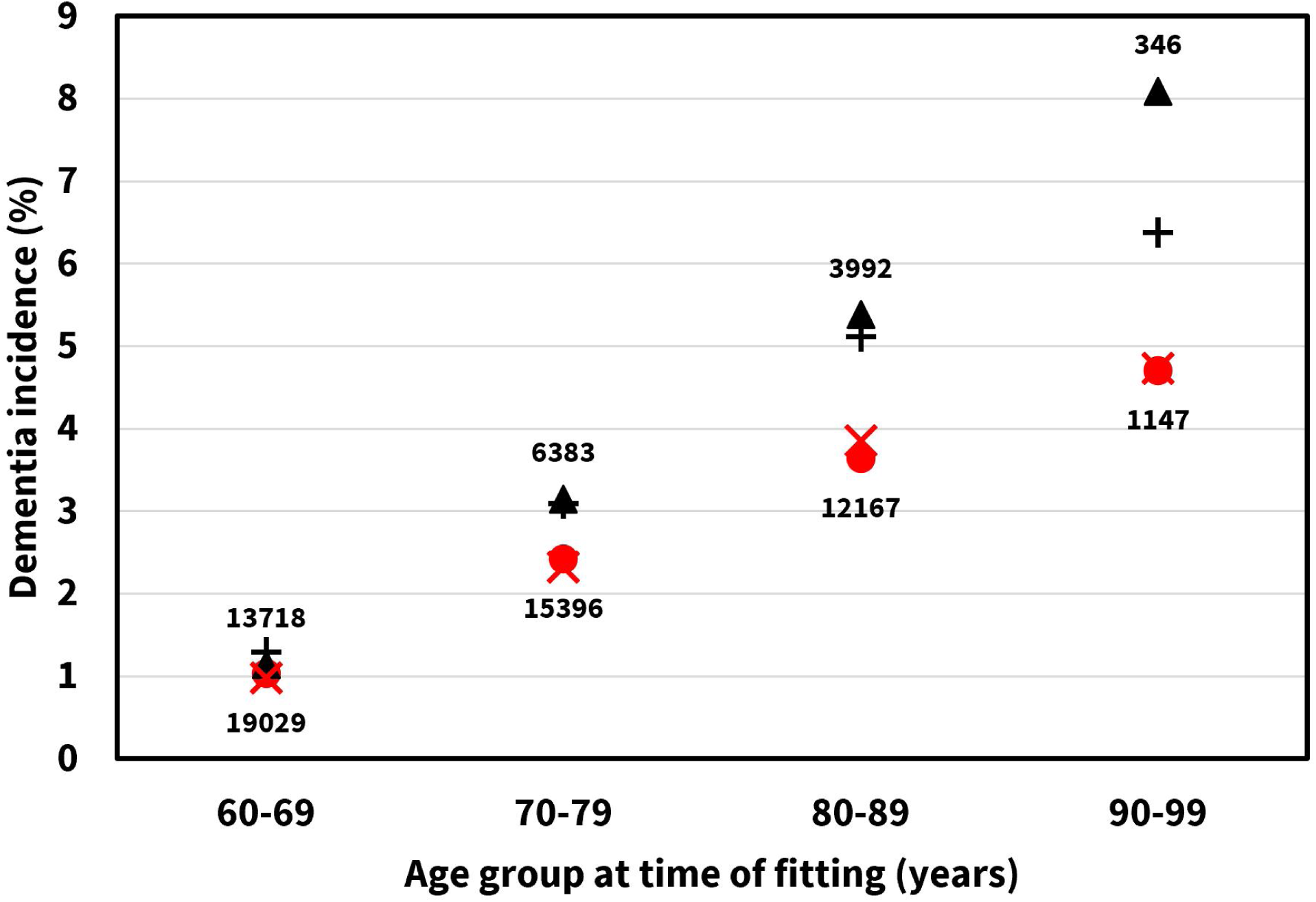
Dementia incidence at 3y6m to 5y post HA fitting for 72,180 patients free of dementia and MCI prior to 3y6m post HA fitting. Filled symbols, unadjusted (raw data); stroke symbols, adjusted (regression model, Table 1; datapoints obtained by assigning each patient their probability of incident dementia predicted by the model, then averaging in each patient age group). Red circle and X, persistent; black triangle and +, non-persistent HA users at 3y6m post fitting. Numbers indicate N for each data point. 2 patients aged 100+ not shown.

As shown, greater HA use persistence is associated with a reduced incidence of dementia, although the effect is marginal for the age group below 70 years (Figure 2). After adjusting for major dementia risk factors, the OR for incident dementia between 3y6m and 5y post HA fitting is 0.73 (95%CI: 0.66 - 0.81) for patients who are persistent HA users at 3y6m, relative to those who are non-persistent.

### Analysis 2: Does cognitive function at time of HA fitting predict subsequent HA use persistence?

Table 2 presents the OR, corresponding 95% CI and p-values for predictors in the logistic regression model of analysis 2. Figure 3 shows the unadjusted and adjusted mean HA use persistence estimates, grouped by cognitive status and age group.

**Table 2:** Logistic regression model for Analysis 2; likelihood of being a persistent hearing-aid user at 3y2m after hearing-aid fitting.

| Predictor | OR (95% CI) | p value |
| --- | --- | --- |
| Age (second-order fit; OR relative to age 75) |  |  |
| 60 years | 0.57 (0.55, 0.58) | <0.001 |
| 70 | 0.88 (0.88, 0.89) | <0.001 |
| 80 | 1.06 (1.05, 1.07) | <0.001 |
| 90 | 0.98 (0.95, 1.00) | 0.10 |
| PTA<sup>a</sup> (per 10 dB increase) | 1.30 (1.30, 1.31) | <0.001 |
| Prevalent-dementia group (vs. high-functioning group) | 0.46 (0.43, 0.48) | <0.001 |
| New hearing-aid recipient (vs. experienced user) | 0.52 (0.51, 0.53) | <0.001 |
| Multimorbidity (per additional body system) | 0.93 (0.92, 0.93) | <0.001 |

<sup>a</sup> PTA; Pure-Tone Average hearing threshold (dB HL).

**Figure 3:**
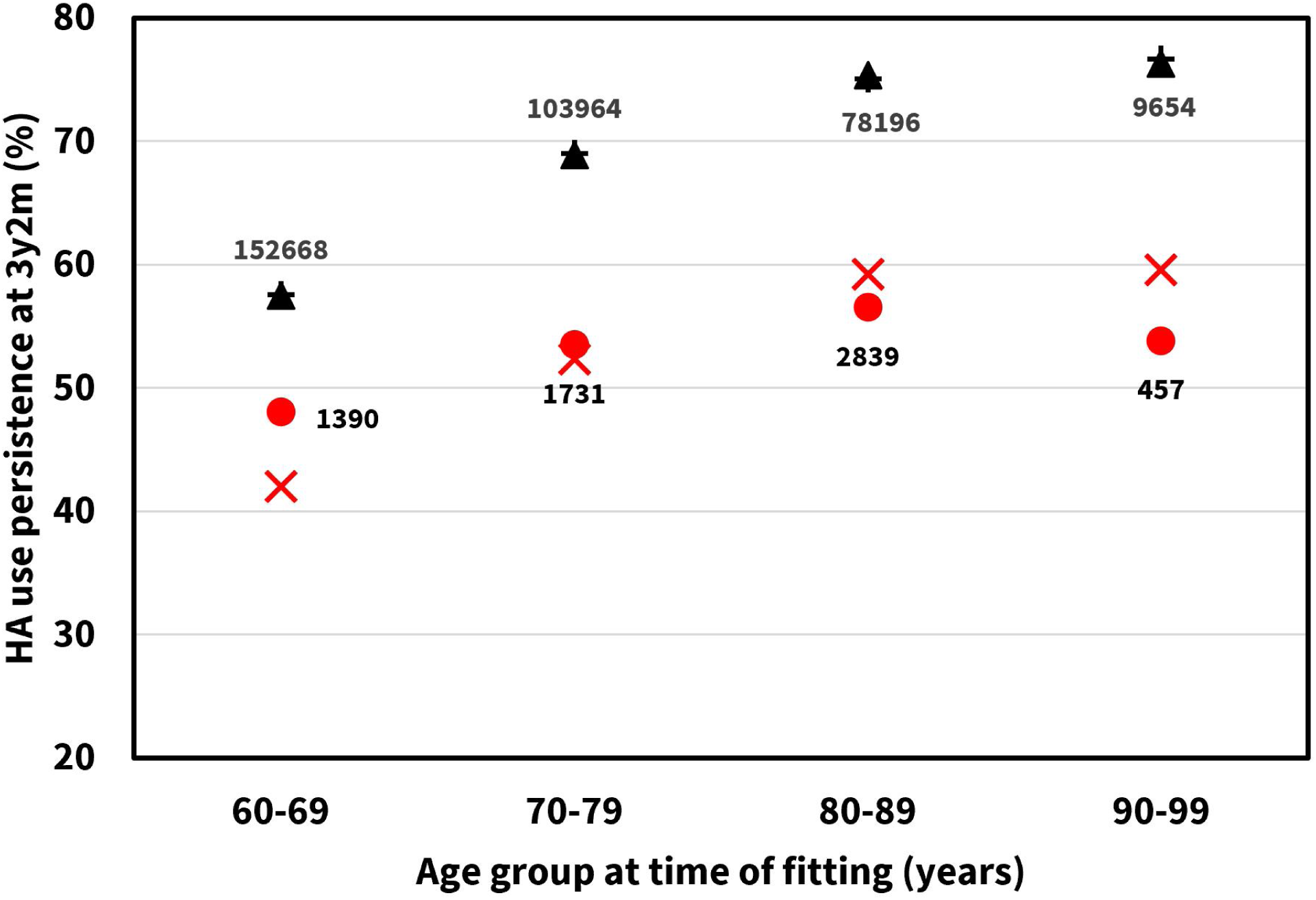
HA use persistence at 3y2m post HA fitting for 350,918 patients. Filled symbols, unadjusted (raw data); stroke symbols, adjusted (regression model, Table 2; datapoints obtained by assigning each patient their probability of HA use persistence predicted by the model, then averaging in each patient age group). Red circle and X, prevalent-dementia group (prevalent dementia at time of HA fitting); black triangle and +, high-functioning group (no dementia or MCI at any time up to 2017/12/31). Numbers indicate N for each data point. 19 patients aged 100+ not shown.

As shown in Figure 3, HA use persistence is about 15 percentage points higher in the normal cognition group compared to the prevalent-dementia group, regardless of age group.

After adjusting for age, multimorbidity, PTA and HA user group (new vs. experienced), the OR for HA use persistence at 3y2m is 0.46 (95%CI: 0.43-0.48) for patients with prevalent dementia at the time of HA fitting, relative to patients with normal cognition (no dementia or MCI diagnosis up to 2017/12/31).

## Discussion

This study provides further evidence that HA use may reduce risk of dementia while also showing that the diagnosis of dementia is associated with lower persistence of HA use.

Consistent with previous findings, we found that patients >60 years old without cognitive impairment at the time of HA fitting, who remained persistent HA users, had 27% reduced odds of receiving a dementia diagnosis 3.5 to 5 years after HA fitting than patients who did not persist in HA use. This suggests that of the four possible mechanisms linking hearing loss and dementia described in the Introduction (Griffiths et al., 2020), the first (common pathology) is not dominant, since HA treatment cannot affect that pathology. Further probing of candidate mechanisms would at the very least require data on duration of hearing loss, which was not available in our dataset.

Patients over 60 years old with a dementia diagnosis prior to HA fitting had 54% reduced odds of being persistent HA users (corresponding to approximately 15 percentage points in each age stratum) compared to those remaining free of MCI and dementia throughout the study time window. This may be due to reduced abilities to perform instrumental activities, as noted in the Introduction (Pérès et al., 2008), or diverse other causes, including memory problems, reduced motivation to engage in social interaction, and carers prioritizing other aspects of care. Our data do not support any distinction between mechanisms on this question.

### Strengths and Limitations

The strengths of this study include a large longitudinal sample, key hearing data, and access to information on a large number of necessary covariates. There are also some limitations. First, the Veteran population is not representative of the general population with respect to health, demographic status or psychosocial characteristics (Gaziano et al., 2015). Hence caution is needed when considering the generalizability of our findings. Nevertheless, there is no reason to think that the underlying mechanisms linking hearing loss and dementia would behave in any fundamentally different ways in a non-Veteran population. Second, one might contend that the care processes in the VA Healthcare System only represent one approach amongst many, and that the effects seen in Analysis 2 are dependent on specific aspects of the VA system. While there might be theoretical merit to this argument, VA care processes in fact vary considerably across a wide network of locations and settings, and are not overly standardized. Furthermore, previous work with the same patient sample (Saunders et al., 2021) indicates that on average, self-reported outcomes from hearing-aid fittings in the VA system are at least as good as those in other healthcare systems. Third, due to the nature of our dataset we had to evaluate HA use persistence at slightly different timepoints in the two analyses, introducing an arbitrary bias. However, analyses on a superset of the present dataset show that HA use persistence is very stable from about 2 years after HA fitting onwards (Zobay et al., 2021). Fourth, by requiring survival to 2017/12/31 we could also have introduced some distortion of the observed effects, since dementia is independently associated with excess mortality (Todd et al., 2013). However, since the dementia- and MCI-free group for analysis 2 was identified on the basis of survival to the end of the study period, not including a survival criterion for other groups would itself have introduced bias.

## Conclusions

This is the first study to combine longitudinal data regarding dementia diagnoses with continuous assessment of HA use in a large patient sample. The results suggest that cognitive impairment is a causative factor in the discontinuation of HA use, while also being consistent with the hypothesis that HA use itself reduces risk of dementia and cognitive decline. This implies that persistent HA use could reduce cognitive decline both in those with, and those without, dementia. However, research studies investigating this must be designed in such a way as to account for the effects of cognitive decline itself on HA use persistence.

In a clinical context, any protective effect of HA use against dementia will not be adequately achieved unless devices and care processes are usable by and accessible to the target population. To encourage and promote persistent HA use, more dementia-friendly devices and care processes are needed.

Finally, from a theoretical perspective, while our data cannot refute the possible existence of a common pathology underlying both hearing loss and dementia, they do not support a strong role for any such pathology.

## Data Availability

All data obtained in the course of this work are the property of the US Dept. of Veterans Affairs. The authors are not authorised to share the data with third parties.

## Acknowledgements

We thank M. Patrick Feeney and ShienPei Silverman for their support throughout the study. Authors GN, LD, OZ and GS made substantial contributions to conception and design, acquisition, analysis and interpretation of data, and drafting and revision of the manuscript. Authors MO and BS made substantial contributions to design, interpretation of data, and critical revision of the manuscript. The views expressed are those of the author(s) and not necessarily those of the VA, NHS, the NIHR or the Department of Health.

## Disclosure statement

The authors declare that they have no competing interests. The funders of this study had no role in the design and conduct of the study; collection, management, analysis, and interpretation of the data; preparation, review, or approval of the manuscript; and decision to submit the manuscript for publication.

## Funding

This work was supported by the Medical Research Council [grant numbers MC_UU_00010/4, MR/S003576/1]; and by the Chief Scientist Office of the Scottish Government; VA Rehabilitation Research and Development grant #9230C, by the VA Office of Academic Affiliations and by the NIHR Manchester Biomedical Research Centre.

## Supplementary material

### Supplementary Data S1: Codesets for dementia, ICD-9 and ICD-10

For diagnosis epochs up to 2015-09-30, the following ICD-9 codes were deemed indicative of dementia primarily associated with age-related endogenous causes:

- 290.XX (dementias)
- 294.1X (dementia in conditions classified elsewhere), 294.2X (dementia, unspecified)
- 331.0 (Alzheimer’s disease), 331.1X (frontotemporal dementia), 331.82 (dementia with Lewy bodies).

For diagnosis epochs from 2015-10-01 onwards, ICD-10 was operative. Domain knowledge and standard translation tools were used to arrive at the following ICD-10 codes, deemed to provide the closest available match to the above ICD-9 set of codes:

- F01.XX (vascular dementia)
- F02.XX (dementia in other diseases classified elsewhere)
- F03.XX (unspecified dementia)
- G30.X (Alzheimer’s disease)
- G31.0X (frontotemporal dementia)
- G31.1 (senile degeneration of brain, not elsewhere classified)
- G31.83 (dementia with Lewy bodies).

### Supplementary Data S2: Codesets for Mild Cognitive Impairment (MCI), ICD-9 and ICD-10

For diagnosis epochs up to 2015-09-30, the following ICD-9 codes was deemed indicative of MCI:

- 331.83 (mild cognitive impairment, so stated)

For diagnosis epochs from 2015-10-01 onwards, the following ICD-10 codes were deemed indicative of MCI:

- G31.84 (mild cognitive impairment, so stated)
- R41.81 (age-related cognitive decline).

### Supplementary Data S3: Codesets for dementia risk factors, ICD-9 and ICD-10

For diagnosis epochs up to 2015-09-30, the following ICD-9 codes were deemed indicative of the given risk factor:

- Obesity: 278.0X (overweight and obesity), 278.1 (localized adiposity).
- Stroke: All codes within 430-438 (cerebrovascular disease).
- Diabetes: All codes within 249 (secondary diabetes mellitus) and 250 (diabetes mellitus).
- Depression: 296.2X (major depressive disorder single episode), 296.3X (major depressive disorder recurrent episode), 296.82 (atypical depressive disorder).
- Bipolar disorder: 296.5X (bipolar I disorder, most recent episode (or current) depressed), 296.6X (bipolar I disorder, most recent episode (or current) depressed), 296.7 (bipolar I disorder, most recent episode (or current) unspecified), 296.8X (other and unspecified bipolar disorders) except for 296.82.
- Hypertension: All codes within 401 (essential hypertension) and 405 (secondary hypertension).

For diagnosis epochs from 2015-10-01 onwards, the following ICD-10 codes were deemed indicative of the given risk factor:

- Obesity: E65 (localized adiposity), E66.XX (overweight and obesity).
- Stroke: All codes in sections I60-I69 (cerebrovascular diseases).
- Diabetes: All codes in sections E08-E13 (diabetes mellitus).
- Depression: All codes in F32 (major depressive disorder, single episode) and F33 (major depressive disorder, recurrent).
- Bipolar disorder: All codes in F31 (bipolar disorder).
- Hypertension: All codes in I10 (essential (primary) hypertension), I15 (secondary hypertension), I16 (hypertensive crisis).

## Notes

### Competing Interest Statement

The authors have declared no competing interest.

## References

Amieva, H., Ouvrard, C., Meillon, C., Rullier, L., & Dartigues, J. F. (2018). Death, depression, disability, and dementia associated with self-reported hearing problems: a 25-year study. The Journals of Gerontology: Series A, 73(10), 1383–1389.

Bertoli, S., Staehelin, K., Zemp, E., Schindler, C., Bodmer, D., & Probst, R. (2009). Survey on hearing aid use and satisfaction in Switzerland and their determinants. International journal of audiology, 48(4), 183–195.

Cohen-Mansfield, J., & Taylor, J. W. (2004). Hearing aid use in nursing homes, Part 2: Barriers to effective utilization of hearing aids. Journal of the American Medical Directors Association, 5(5), 289–296.

Dawes, P., Cruickshanks, K. J., Fischer, M. E., Klein, B. E., Klein, R., & Nondahl, D. M. (2015). Hearing-aid use and long-term health outcomes: Hearing handicap, mental health, social engagement, cognitive function, physical health, and mortality. International journal of audiology, 54(11), 838–844.

Deal, J. A., Sharrett, A. R., Albert, M. S., Coresh, J., Mosley, T. H., Knopman, D., Wruck, L.M., & Lin, F. R. (2015). Hearing impairment and cognitive decline: a pilot study conducted within the atherosclerosis risk in communities neurocognitive study. American journal of epidemiology, 181(9), 680–690.

Fischer, M. E., Cruickshanks, K. J., Wiley, T. L., Klein, B. E., Klein, R., & Tweed, T. S. (2011). Determinants of hearing aid acquisition in older adults. American journal of public health, 101(8), 1449–1455.

Gaziano, J. M., Concato, J., Galea, S., Smith, N. L., & Provenzale, D. (2015). Epidemiologic approaches to veterans’ health. Epidemiologic reviews, 37(1), 1–6.

Gopinath, B., Schneider, J., Hartley, D., Teber, E., McMahon, C. M., Leeder, S. R., & Mitchell, P. (2011). Incidence and predictors of hearing aid use and ownership among older adults with hearing loss. Annals of epidemiology, 21(7), 497–506.

Gregory, S., Billings, J., Wilson, D., Livingston, G., Schilder, A. G., & Costafreda, S. G. (2020). Experiences of hearing aid use among patients with mild cognitive impairment and Alzheimer’s disease dementia: A qualitative study. SAGE open medicine, 8, 2050312120904572.

Griffiths, T.D., Lad, M., Kumar, S., Holmes, E., McMurray, B., Maguire, E.A., Billig, A.J. & Sedley, W. (2020). How can hearing loss cause dementia?. Neuron. https://doi.org/10.1016/j.neuron.2020.08.003.

Healthcare Cost and Utilization Project (HCUP). HCUP Chronic Condition Indicator. May 2016. Agency for Healthcare Research and Quality, Rockville, MD. Available at: www.hcup-us.ahrq.gov/toolssoftware/chronic/chronic.jsp. Accessed December 1, 2020.

Livingston, G., Huntley, J., Sommerlad, A., Ames, D., Ballard, C., Banerjee, S., … Mukadam, N. (2020). Dementia prevention, intervention, and care: 2020 report of the Lancet Commission. The Lancet, 396(10248), 413–446.

Maharani, A., Dawes, P., Nazroo, J., Tampubolon, G., Pendleton, N., SENSE-Cog WP1 group, … & von Hanno, T. (2018). Longitudinal relationship between hearing aid use and cognitive function in older Americans. Journal of the American Geriatrics Society, 66(6), 1130–1136.

Mahmoudi, E., Basu, T., Langa, K., McKee, M. M., Zazove, P., Alexander, N., & Kamdar, N. (2019). Can hearing aids delay time to diagnosis of dementia, depression, or falls in older adults?. Journal of the American Geriatrics Society, 67(11), 2362–2369.

Pérès, K., Helmer, C., Amieva, H., Orgogozo, J. M., Rouch, I., Dartigues, J. F., & Barberger-Gateau, P. (2008). Natural history of decline in instrumental activities of daily living performance over the 10 years preceding the clinical diagnosis of dementia: a prospective population-based study. Journal of the American Geriatrics Society, 56(1), 37–44.

Ray, J., Popli, G., & Fell, G. (2018). Association of cognition and age-related hearing impairment in the English longitudinal study of ageing. JAMA Otolaryngology– Head & Neck Surgery, 144(10), 876–882.

Saunders, G. H., Dillard, L. K., Zobay, O., Cannon, J. B., & Naylor, G. (2021). Electronic Health Records As a Platform for Audiological Research: Data Validity, Patient Characteristics, and Hearing-Aid Use Persistence Among 731,213 US Veterans. Ear and hearing, 42(4), 927–940.

Todd, S., Barr, S., Roberts, M., & Passmore, A. P. (2013). Survival in dementia and predictors of mortality: a review. International journal of geriatric psychiatry, 28(11), 1109–1124.

Zobay, O., Dillard, L. K., Naylor, G., & Saunders, G. H. (2021). A measure of long-term hearing-aid use persistence based on battery re-ordering data. Ear and hearing. doi: 10.1097/AUD.0000000000001032

